# Clinically aligned rationale generation for glaucoma subtype classification via a knowledge-distilled language model

**DOI:** 10.64898/2026.06.15.26355522

**Authors:** Mousa Moradi, Asahi Fujita, Niloufar Bineshfar, Daniel M. Vu, Kanza Aziz, Daniel L. Liebman, Mengyu Wang, Tobias Elze, Mohammad Eslami, Nazlee Zebardast

## Abstract

Automated glaucoma subtype classification from clinical notes remains clinically unactionable without subspecialty-aligned explanations supporting clinician-facing deployment. We extended our Ci-SSGAN with a GPT-5.2-to-Qwen3-8B teacher-distilled reasoning module, fine-tuning Qwen3-8B on 2,660 de-identified ophthalmology notes using expert-reviewed rationales. On 294 notes, the fine-tuned model achieved ROUGE-L 0.792 ± 0.013 and BERTScore F1 0.955 ± 0.004, surpassing eight zero-shot comparators including GPT-4o and GPT-4.1, establishing privacy-preserving distillation as a path to interpretable AI.

## Main Text

Artificial intelligence applied to free-text electronic health records (EHRs) is reshaping disease characterization, risk stratification, and clinical decision support across medicine ^1–3^. Clinical notes capture diagnostic reasoning, phenotypic nuance, and longitudinal disease context that structured billing codes including ICD-10 assignments, systematically fail to represent ^4,5^. This limitation is particularly consequential in ophthalmology: glaucoma, the leading cause of irreversible blindness worldwide and projected to affect 111.8 million people by 2040 ^6^, comprises mechanistically and therapeutically distinct subtypes whose ICD-10 coding achieves only ~81% accuracy and less than 50% specificity for subtype discrimination ^7,8^. Clinical notes from glaucoma specialist encounters contain the phenotypic evidence (e.g., gonioscopy findings, disc morphology, intraocular pressure patterns, assessment of structural and functional visual changes) required to resolve these distinctions but remain largely inaccessible to automated systems without specialized annotation frameworks ^9^. Our previously published Ci-SSGAN framework addressed the annotation challenge by combining 2,954 expert-curated notes with 349,587 unlabeled ophthalmology records in a demographically conditioned semi-supervised generative adversarial network, achieving 0.949 AUROC and substantially narrowing subgroup performance gaps relative to ICD-based baselines ^10^.

Yet classification alone is not sufficient for trustworthy clinical AI ^11–14^. In practice, clinicians must understand not only what subtype is predicted, but why a note supports that prediction. This need is particularly important in glaucoma, where subtle distinctions among open-angle, angle-closure, exfoliative, pigmentary, and other secondary mechanisms depend on specific textual evidence embedded in assessment and plan narratives. Recent large language models have shown strong capabilities in clinical text processing and medical reasoning ^15,16^, but off-the-shelf zero-shot systems remain inconsistent for subspecialty note interpretation, and they do not necessarily produce explanations aligned with expert ophthalmic reasoning.

Building directly on Ci-SSGAN’s subtype predictions ^10^, we employed a generative knowledge distillation strategy ^17,18^ in which GPT-5.2 (accessed through the Mass General Brigham AI Zone HIPAA-compliant environment on Microsoft Azure AI Foundry) served as the teacher model, generating one-sentence diagnostic rationales conditioned on each note and its Ci-SSGAN-assigned subtype label — explaining why that predicted subtype is clinically supported by the note text. All teacher outputs underwent mandatory expert review by a fellowship-trained glaucoma specialist prior to inclusion as supervision targets, ensuring that only clinically validated rationales were used for student model training. To further constrain hallucination at inference, the system prompt explicitly instructed both the teacher and student models not to fabricate clinical findings absent from the source note, enforced a fixed one-sentence output format anchored to observable note content, and required all rationales to conclude with a subtype drawn exclusively from the predefined priority hierarchy (full prompt in Supplementary Table S1). The student model (Qwen3-8B) was fine-tuned locally within MGB-controlled infrastructure using these expert-reviewed rationales via QLoRA ^19^, a parameter-efficient adaptation strategy that enables full-scale language model fine-tuning under memory constraints ^20^. At inference, Ci-SSGAN assigns the glaucoma subtype and the fine-tuned Qwen3-8B module generates the corresponding rationale, coupling classification and interpretation into a single auditable output (Fig. 3B).

The fine-tuned Qwen3-8B achieved overall ROUGE-1 of 0.8033 ± 0.011, ROUGE-L of 0.7921 ± 0.013, BERTScore F1 of 0.9545 ± 0.004, and LLM-judge score of 4.56 ± 0.06 out of 5 on the held-out test set of 294 notes from 105 patients (Table 1, Fig. 1A). The fine-tuned model significantly outperformed all eight zero-shot comparators across all four metrics (*P* value < 0.001 for all). Relative to its zero-shot counterpart, fine-tuning yielded 151.6% improvement in ROUGE-1 (0.3194 vs. 0.8033), 213.5% improvement in ROUGE-L (0.2527 vs. 0.7921), 8.4% improvement in BERTScore F1 (0.8802 vs. 0.9545), and 20.0% improvement in LLM-judge score (3.80 vs. 4.56/5), all *P* value < 0.001. Among the strongest zero-shot comparators, GPT-4o achieved ROUGE-1 and ROUGE-L scores 50.7% and 54.4% lower than the fine-tuned model, respectively, with BERTScore F1 26.8% lower. GPT-4.1 attained an LLM-judge score of 4.46/5, 0.10 points below the fine-tuned model, with ROUGE-L 53.7% lower. GPT-OSS-120B, despite its substantially larger parameter count, achieved BERTScore F1 of 0.8650 (9.5% below the fine-tuned Qwen3-8B) with ROUGE-L 72.7% lower and an LLM-judge score of 3.96/5, 13.2% below the fine-tuned model. MedGemma-1.5-4B and LLaMA-3.2-3B, both domain-relevant medical models, fell 67.9% and 74.5% below the fine-tuned model in ROUGE-L, respectively. DeepSeek variants performed markedly worse across lexical and semantic metrics, with DeepSeek-LLaMA-8B achieving ROUGE-L 84.8% and BERTScore F1 54.4% below the fine-tuned model, and LLM-judge scores of 2.21/5 and 2.01/5 for DeepSeek-Qwen-7B and DeepSeek-LLaMA-8B respectively, reflecting poor alignment with expert clinical reasoning.

**Table 1.** Overall rationale-generation performance across compared models. NA indicates that an LLM-judge score was not reported for that model. ZS= Zero-shot; FT= Fine-tuned. GPT-4o and GPT-4.1 were accessed through MGB AI Zone. NA= Not Applicable.

| Model | ROUGE-1 | ROUGE-L | BERTScore F1 | LLM-Judge (/5) |
| --- | --- | --- | --- | --- |
| Qwen3-8B (ZS)<sup>21</sup> | 0.3194 | 0.2527 | 0.8802 | 3.80 |
| **Qwen3-8B (FT)** | **0.8033** | **0.7921** | **0.9545** | **4.56** |
| MedGemma-1.5-4B<sup>22</sup> | 0.3181 | 0.2539 | 0.6965 | 3.99 |
| LLaMA-3.2-3B<sup>23</sup> | 0.2554 | 0.2022 | 0.6685 | 3.90 |
| GPT-4o | 0.3960 | 0.3609 | 0.6993 | NA |
| GPT-4.1 | 0.3721 | 0.3037 | 0.7101 | 4.46 |
| GPT-OSS-120B<sup>24</sup> | 0.2769 | 0.2161 | 0.8650 | 3.96 |
| DeepSeek-Qwen-7B<sup>25</sup> | 0.2089 | 0.1721 | 0.6310 | 2.21 |
| DeepSeek-LLaMA-8B<sup>25</sup> | 0.1508 | 0.1202 | 0.4357 | 2.01 |

**Fig. 1.**
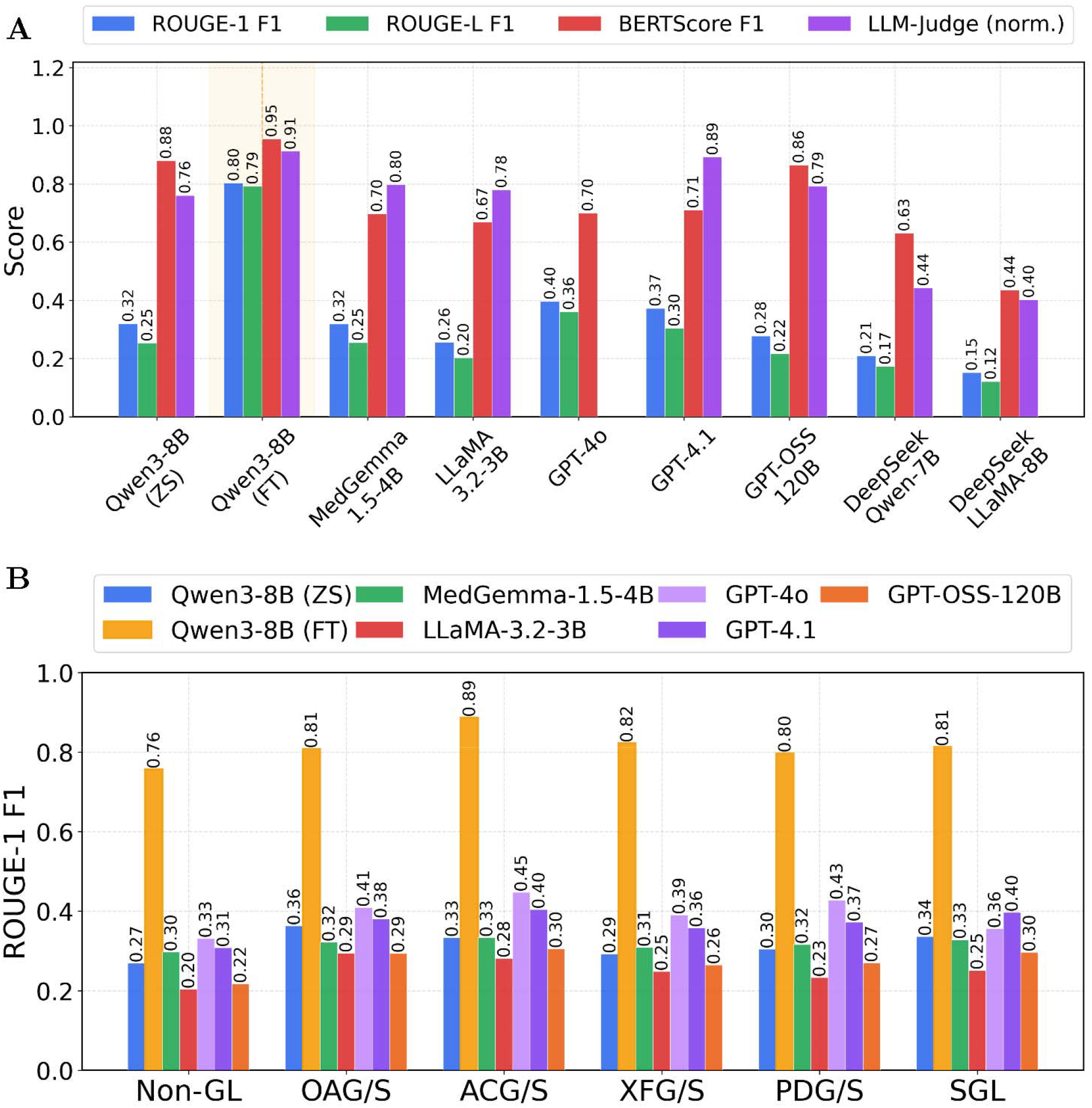
Reasoning quality benchmarks across nine models. (A) Overall ROUGE-1, ROUGE-L, BERTScore F1, and normalized LLM-Judge scores; fine-tuned Qwen3-8B (highlighted) leads all metrics. (B) Per-class ROUGE-1 F1 across six glaucoma subtypes, with fine-tuned Qwen3-8B consistently outperforming all zero-shot baselines by a wide margin.

Per-class ROUGE-1 F1 for the fine-tuned Qwen3-8B ranged from 0.757 to 0.887 across the six glaucoma subtypes, representing 2.1- to 2.9-fold improvements over zero-shot Qwen3-8B, with values of 0.757 for non-glaucoma, 0.808 for OAG/S, 0.887 for ACG/S, 0.823 for XFG/S, 0.797 for PDG/S, and 0.813 for SGL (Fig. 1B). Per-class gains over zero-shot Qwen3-8B were significant for all six subtypes (*P* value < 0.001), with the largest relative improvement in ACG/S (0.334 vs. 0.887; +165.6%) and the smallest in OAG/S (0.363 vs. 0.808; +122.6%). Against the strongest zero-shot lexical comparator, GPT-4o, the fine-tuned model exceeded per-class ROUGE-1 by 98.5% for non-glaucoma, 97.3% for OAG/S, 98.4% for ACG/S, 110.5% for XFG/S, 86.9% for PDG/S, and 128.4% for SGL, reflecting consistent gains across both common and rare subtypes. The fine-tuned model significantly outperformed all remaining zero-shot comparators on per-class ROUGE-1 across all subtypes (*P* value < 0.001 for all).

Per-class LLM-judge scores for the fine-tuned Qwen3-8B ranged from 4.24 to 4.73 out of 5, reflecting uniformly high and consistent qualitative ratings across all six subtypes (Fig. 2A). Compared with zero-shot Qwen3-8B, the fine-tuned model achieved clinically meaningful gains of +0.99 points for OAG/S (+29.3%), +0.84 for ACG/S (+21.9%), +0.94 for XFG/S (+25.1%), +0.90 for PDG/S (+24.4%), and +0.64 for SGL (+15.6%), all *P* value < 0.001, with no significant difference for non-glaucoma (4.24 vs. 4.29; P = 0.61), where both models demonstrated similarly strong performance. GPT-4.1 achieved per-class LLM-judge scores of 4.19 (Non-GL), 4.30 (OAG/S), 4.66 (ACG/S), 4.47 (XFG/S), 4.48 (PDG/S), and 4.67 (SGL), falling below the fine-tuned model on all six subtypes by margins of 0.02 to 0.22 points. GPT-OSS-120B achieved per-class scores ranging from 3.87 to 4.05, all significantly below the fine-tuned model (*P* value < 0.001). Per-class BERTScore F1 for the fine-tuned Qwen3-8B ranged from 0.938 to 0.969, representing 5.4% to 10.9% absolute improvements over zero-shot Qwen3-8B across subtypes (*P* value < 0.001 for all six) and 8.1% to 16.3% improvements over the next-best zero-shot BERTScore comparator, GPT-OSS-120B, across subtypes (*P* value < 0.001) (Fig. 2B). The ACG/S subtype achieved the highest fine-tuned BERTScore F1 at 0.969, representing a 10.9% gain over zero-shot Qwen3-8B and a 12.7% gain over GPT-OSS-120B. DeepSeek-LLaMA-8B achieved per-class BERTScore F1 values ranging from 0.364 to 0.517, falling 46.8% to 62.4% below the fine-tuned model across subtypes.

**Fig. 2.**
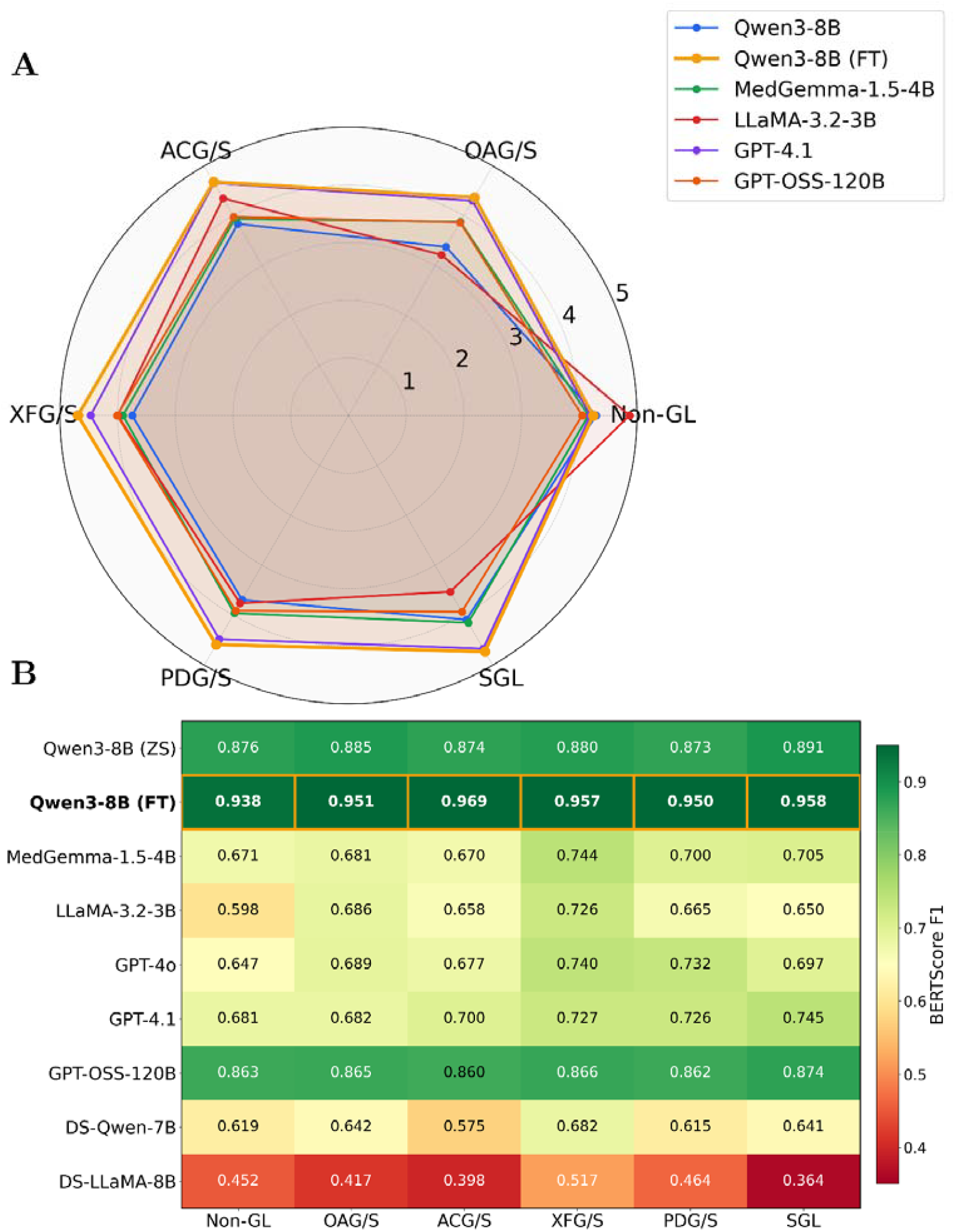
Per-class semantic reasoning quality across models. (A) Radar chart of LLM-Judge scores (1–5) across six glaucoma subtypes; fine-tuned Qwen3-8B (orange) achieves the broadest and most uniform coverage. (B) BERTScore F1 heatmap by subtype, with fine-tuned Qwen3-8B (highlighted row) attaining the highest semantic similarity to expert rationales across all six classes (0.938–0.969).

The diagnosis-conditioned keyword activation map revealed that the fine-tuned Qwen3-8B acquired distinct, subtype-specific lexical fingerprints rather than relying on generic clinical language (Fig. 3A). Among the top 30 globally selected keywords, four showed strong class-exclusive activation with gaps exceeding 0.35 above the second-highest class: “pseudoexfoliation” for XFG/S (activation score 0.89, gap of 0.39 above ACG/S), “secondary” for SGL (0.66, gap of 0.47 above OAG/S), “closure” for ACG/S (0.76, gap of 0.43 above OAG/S), and “syndrome” for XFG/S (0.50, gap of 0.38 above ACG/S). Template-level keywords shared across all six subtypes (“glaucoma,” “diagnosis,” “priority,” and “mentions”) showed uniformly high activation scores (0.44–1.00), reflecting the structured one-sentence rationale format enforced during fine-tuning. Non-glaucoma and OAG/S exhibited broader, less discriminative activation profiles, consistent with their clinically overlapping and exclusion-based definitions, whereas ACG/S, XFG/S, PDG/S, and SGL showed mechanistically specific keywords with elevated activation. Consistent with these patterns, end-to-end inference demonstrates that the model generates subtype-consistent rationales grounded in key clinical findings (Fig. 3B). Classification and reasoning outputs for 6 randomly selected patients with 30 total notes in our test set (294 notes from 105 patients) are provided in Supplementary Table S2. At the note level, Ci-SSGAN assigns the highest-probability class as the predicted subtype; when confidence exceeds 70%, the reasoning module generates a single-sentence rationale grounded exclusively in that class and sets Human Review to False, whereas at confidence ≤70%, a two-sentence rationale is generated covering both the top and second-highest probability classes with supporting clinical evidence for each, and Human Review is set to True, flagging the note for specialist oversight. At the patient level, where longitudinal records may yield multiple discordant note-level predictions, a single definitive subtype is assigned by applying the clinical priority hierarchy (XFG/S > PDG/S > SGL > ACG/S > OAG/S > Non-GL) across all notes for a given patient, encoding the etiological specificity and irreversibility of higher-priority diagnoses: once exfoliation syndrome, pigment dispersion, or a secondary mechanism is documented in any note in the longitudinal record, it supersedes lower-priority predictions regardless of note frequency.

**Fig. 3.**
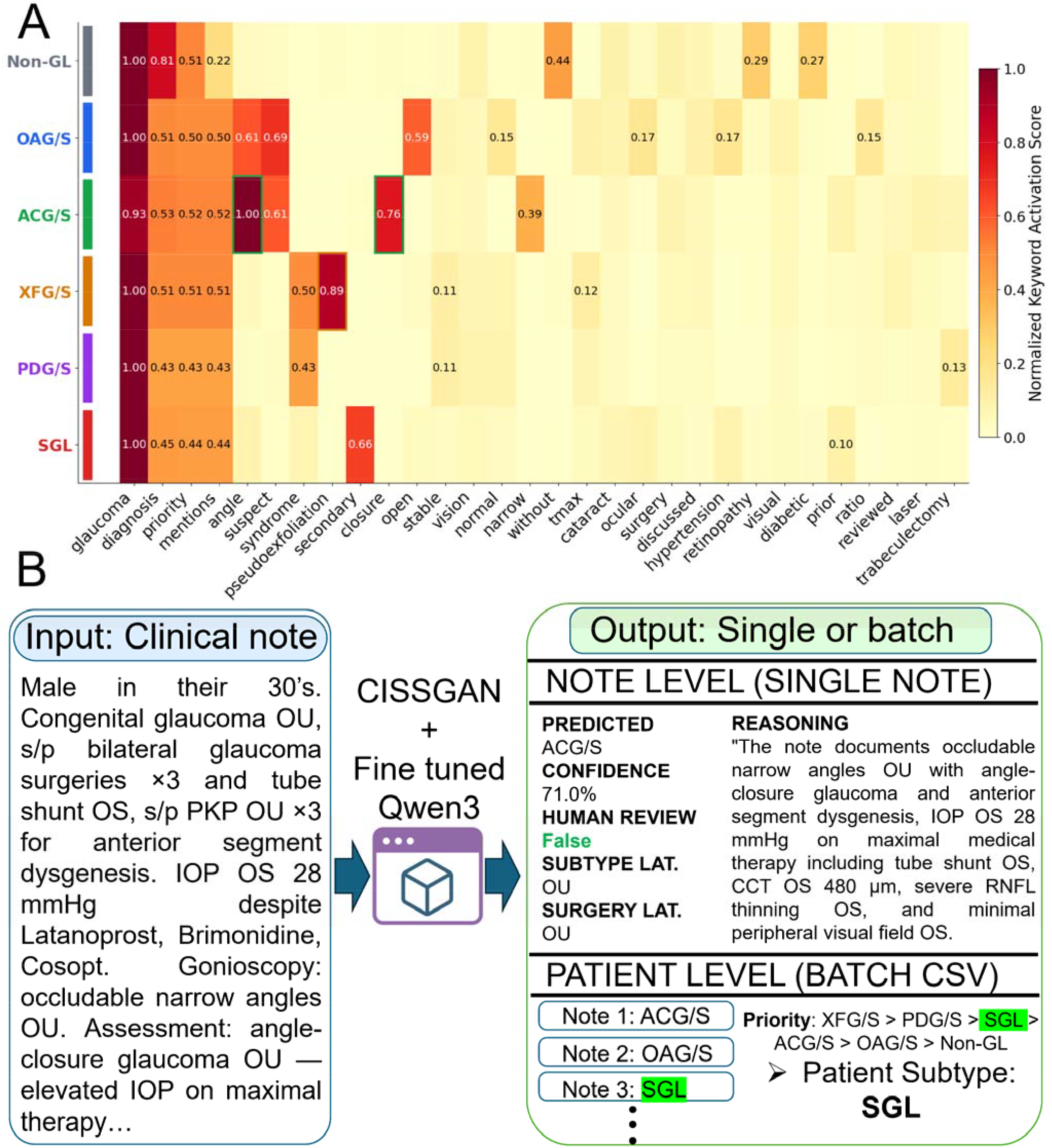
Clinically aligned keyword activation and end-to-end reasoning generation. (A) Diagnosis-conditioned keyword activation map for the fine-tuned Qwen3-8B model, showing normalized activation of the top 30 clinical keywords across six glaucoma subtypes. Class-specific signatures are evident (e.g., “pseudoexfoliation” for XFG/S, “closure” for ACG/S, “suspect” for OAG/S). Highlighted cells indicate high class specificity (activation gap >0.35). (B) Example end-to-end inference via the publicly accessible Hugging Face demonstration interface. A de-identified clinical note is processed to produce subtype classification (OAG/S, confidence 99.7%), laterality, and a concise subtype-consistent rationale. At the note level, predictions with confidence >70% bypass human review and receive a single-sentence rationale based on the top predicted class; predictions with confidence ≤70% are flagged for specialist review, with a two-sentence rationale summarizing the top two predicted classes and their supporting clinical evidence. At the patient level, a single definitive subtype is assigned using the clinical priority hierarchy: XFG/S > PDG/S > SGL > ACG/S > OAG/S > Non-GL.

This study demonstrates that generative knowledge distillation — in which a large teacher model provides target text sequences rather than soft logits — enables a compact, locally deployable language model fine-tuned on a domain-specific glaucoma corpus to produce rationale quality substantially superior to frontier zero-shot systems, including GPT-4o and GPT-4.1, on both lexical and semantic metrics. The finding that task-specific adaptation outperforms larger general-purpose models challenges the assumption that scale alone is sufficient for subspecialty clinical reasoning and establishes parameter-efficient generative knowledge distillation as a viable, privacy-preserving strategy for embedding interpretability into clinical AI workflows. The reasoning module is architecturally modular and can be directly coupled with live Ci-SSGAN subtype predictions, enabling clinicians to review not only the predicted glaucoma subtype but also the supporting rationale as an auditable output. Consistent with the future directions outlined in the original Ci-SSGAN publication, subsequent work should extend this framework toward multimodal reasoning that integrates optical coherence tomography, visual field indices, and structured EHR data alongside free-text notes, evaluate the reasoning module prospectively in terms of clinician trust, workflow integration, and diagnostic utility, and validate generalizability across external institutions with demographically distinct patient populations.

Limitations include the single-center origin of the dataset, which constrains generalizability across institutions, note styles, and demographic distributions. LLM-judge evaluation was not performed for GPT-4o due to API access constraints, representing a minor gap in the comparative semantic evaluation. Additionally, the LLM-judge evaluation framework, while widely adopted, introduces subjectivity dependent on the judge model’s own clinical alignment, and prospective human evaluation by fellowship-trained glaucoma specialists on model-generated rationales remains necessary before clinical deployment.

The proposed classification and reasoning framework enables clinically aligned, auditable rationale generation alongside automated glaucoma subtype classification, supporting clinician trust and facilitating integration of AI-assisted interpretation into ophthalmology workflows.

## Methods

### Cohort selection, de-identification, and data preprocessing

The expert-curated glaucoma note cohort was drawn from the MEE EHR system, which encompasses over 250 million clinical notes across all departments; ophthalmology notes constitute 3.2% of the total, with physician-generated progress notes and assessment-and-plan notes from May 2015 through December 2024 comprising the primary source pool. This study reused the expert-curated glaucoma note cohort described in the original Ci-SSGAN publication ^10^. Briefly, the cohort comprised 2,954 notes from 1,105 patients across six diagnostic categories (Non-GL, OAG/S, ACG/S, XFG/S, PDG/S, SGL), adjudicated by six fellowship-trained glaucoma specialists (Cohen’s kappa = 0.90). The dataset was partitioned at the patient level into a 90% development set (2,660 notes from 1,000 patients) and a 10% held-out test set (294 notes from 105 patients), with five-fold cross-validation performed within the development partition. The identical split from the original study was preserved to ensure direct comparability, with no patient overlap between training and test sets at any stage. All notes were de-identified using the Philter package ^26^, and processed with a 64-token overlap sliding window strategy for length-aware context preservation, following the original preprocessing pipeline ^10^. Teacher rationales were generated for the 2,660-note development set as discrete target text sequences for sequence-level supervision, using GPT-5.2 through the Mass General Brigham (MGB) AI Zone HIPAA-compliant environment, with each note and assigned label provided via a structured prompt requiring exactly one concise clinical sentence grounded in the note text and consistent with the established subtype priority hierarchy (XFG/S > PDG/S > SGL > ACG/S > OAG/S > Non-GL). All rationales were reviewed and approved by a fellowship-trained glaucoma specialist prior to use as supervision targets. Ethical approval was obtained from the MGB Institutional Review Board under the same protocol as the original study, with waiver of informed consent.

### Model development and architecture

Fine-tuning was performed using a QLoRA framework ^19^ on Qwen3-8B ^21^. The base model was loaded in 4-bit NF4 with double quantization to reduce memory while preserving performance. LoRA ^20^ was applied to query, key, value, output, gate, up, and down projections (rank 32, alpha 64, dropout 0.1, no bias), restricting updates to adapter parameters while freezing base weights (Fig. 4A). Training used a structured chat template with a system message defining clinical role, diagnostic rules, and output format; a user message containing a de-identified note (≤1,200 characters) and assigned label; and an assistant message with the gold-standard rationale. To minimize hallucination, the system prompt explicitly instructed the model not to fabricate clinical findings absent from the source note, constrained output to a single sentence anchored in observable note content, and enforced a fixed subtype priority vocabulary for the concluding clause, ensuring generated rationales remained traceable to source text rather than model priors (full system prompt provided in Supplementary Table S1). Completion-only training was enforced by computing loss exclusively over generated rationale tokens.

**Fig. 4.**
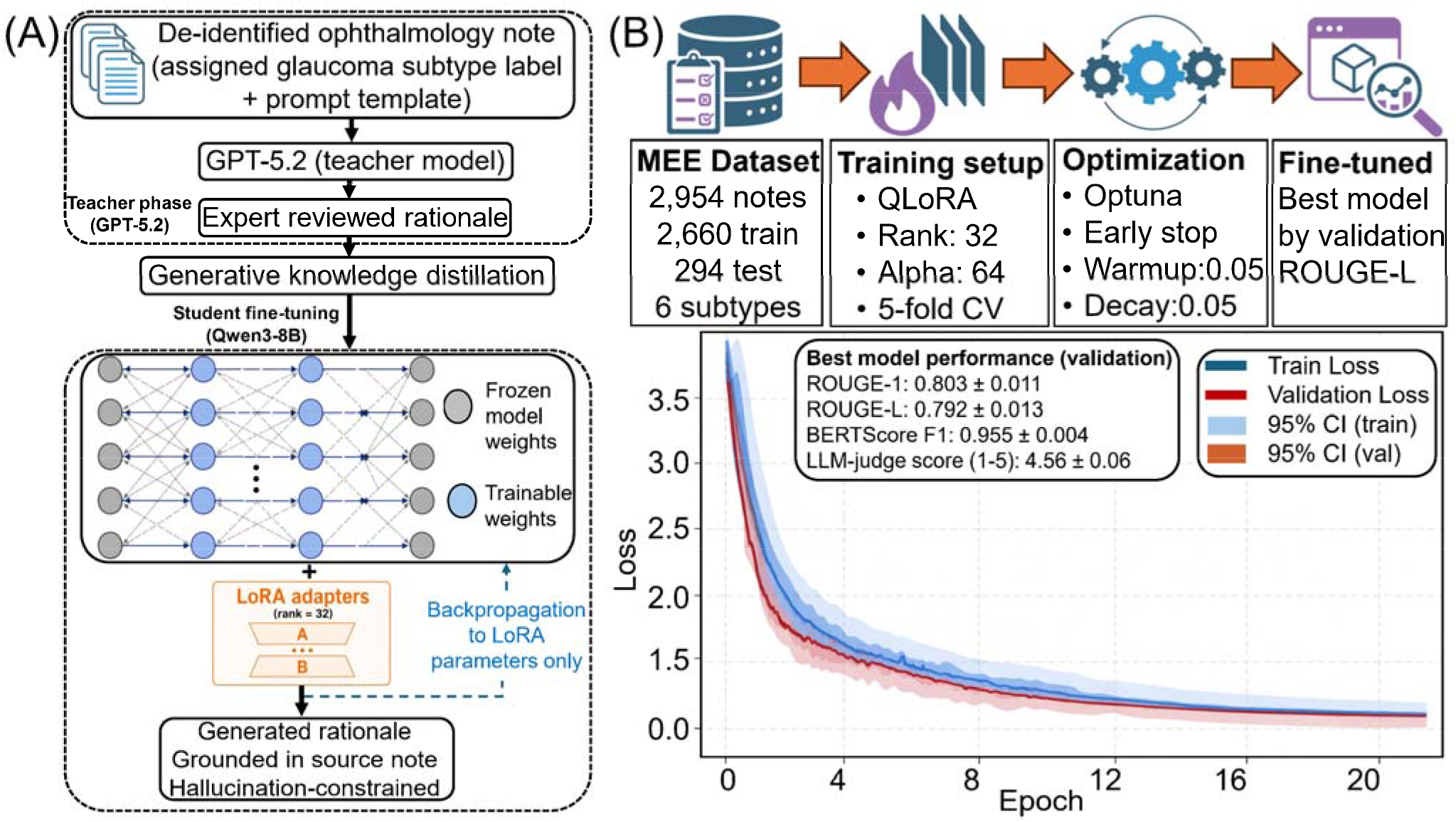
QLoRA fine-tuning pipeline and training dynamics for glaucoma rationale generation. (A) Qwen3-8B is fine-tuned in 4-bit NF4 precision using low-rank LoRA adapters (rank 32), with frozen base weights and backpropagation restricted to adapter parameters. Inputs consist of de-identified ophthalmology notes, assigned glaucoma subtype labels, and a structured prompt template; outputs are expert-supervised one-sentence rationales. (B) Training setup (2,660 notes, 5-fold CV, QLoRA) and loss curves averaged across folds, showing rapid convergence by epoch 8 with no overlap between train and validation loss. Best validation checkpoint achieved ROUGE-L 0.792 ± 0.013 and LLM-judge score 4.56 ± 0.06.

Five-fold patient-level cross-validation was used with maximum sequence length 2,048, generation cap 60 tokens, batch size 2 with gradient accumulation (effective batch size 8), learning rate *1 × 10*^−5^, cosine scheduling, 0.05 warmup ratio and weight decay, gradient checkpointing, and paged AdamW 8-bit optimization. Models were trained up to 100 epochs with early stopping (patience 7) based on validation ROUGE-L, evaluated each epoch. Hyperparameters were tuned using Optuna ^27^ (50 trials). Best checkpoints were selected per fold by validation ROUGE-L. Training was conducted on a single NVIDIA H100 (80GB) using PyTorch 2.3 (bfloat16, TF32). Convergence occurred by epoch 8 with aligned train/validation loss and no overfitting (Fig. 4B). Final performance was evaluated on a held-out test set (n=294), reported as mean ± standard deviation across folds.

Zero-shot comparators included Qwen3-8B ^21^, MedGemma-1.5-4B ^22^, LLaMA-3.2-3B ^23^, GPT-4o, GPT-4.1, GPT-OSS-120B ^24^, DeepSeek-Qwen-7B ^25^, and DeepSeek-LLaMA-8B ^25^, all using identical structured prompts without fine-tuning. GPT-4o and GPT-4.1 were accessed via the MGB AI Zone; all others were deployed locally. Fine-tuned adapters and quantized checkpoints are publicly available on Hugging Face for reproducibility and downstream use.

### Model explainability

To characterize the clinical vocabulary learned by the fine-tuned Qwen3-8B during teacher-distilled training, we constructed a diagnosis-conditioned keyword activation map from model-generated rationales on the held-out test set. For each note in the test set, the fine-tuned model generated a one-sentence rationale under greedy decoding with repetition penalty 1.1 and a maximum of 60 new tokens, with thinking mode disabled. Keywords were extracted from generated rationales using regular expression tokenization, and per-class keyword frequency was weighted by a factor of 2.0 to reflect the high informational content of generated outputs relative to source note terms, which were additionally extracted at a weight of 0.3 to capture contextual clinical signal. Stopwords, tokens shorter than three characters, non-alphabetic tokens, and tokens appearing in fewer than two classes above a frequency threshold of 1.0 were excluded. The top 30 globally ranked keywords were selected based on aggregate weighted frequency across all six classes, and a normalized per-class activation matrix was constructed by dividing each class row by its maximum value. Class-specific keywords were identified as those with a normalized activation gap exceeding 0.35 above the second-highest class, and their positions were annotated with bordered highlights in the heatmap visualization (Fig. 3). Keyword activation data were additionally exported in JSON format for supplementary inspection.

### Inference deployment and decision logic

At the note level, predicted subtype and confidence were derived directly from Ci-SSGAN softmax probabilities, with the highest-probability class assigned as the note-level subtype. When classifier confidence exceeded 70%, the reasoning module generated a single-sentence rationale grounded exclusively in the top predicted class using specific clinical evidence from the note (named syndromes, medications, surgical history, intraocular pressure values, visual field findings, or structural diagnoses), and Human Review was set to False. When confidence was ≤70%, a two-sentence rationale was generated: the first sentence justified the top predicted class with supporting clinical evidence, and the second explained why the second-highest probability class also received classifier signal; Human Review was set to True, flagging the note for specialist oversight. All six class probability scores were returned alongside the predicted subtype to support clinical transparency and auditable review. At patient level, where multiple longitudinal notes may yield discordant note-level subtype predictions, a single definitive subtype was assigned by applying a clinical priority hierarchy (XFG/S > PDG/S > SGL > ACG/S > OAG/S > Non-GL) across all notes for a given patient. This hierarchy encodes the etiological specificity and diagnostic irreversibility of higher-priority subtypes: once exfoliation syndrome, pigment dispersion, or a secondary mechanism is documented in any note in the longitudinal record, it supersedes lower-priority predictions regardless of note frequency. Note-level outputs (six-class probability distribution, predicted subtype, subtype and surgery laterality, confidence, Human Review flag, and reasoning) and patient-level outputs (priority-assigned definitive subtype, mean confidence, note count) were exported as separate comma-separated value files to support downstream clinical review and research analysis.

### Metrics and statistical analysis

Rationale quality was assessed using ROUGE-1 and ROUGE-L F1 ^28^ (rouge-score library, stemming enabled), BERTScore F1 ^29^ (microsoft/deberta-xlarge-mnli checkpoint), and LLM-judge scoring ^30^ on a 1–5 scale evaluating clinical accuracy, specificity, output format adherence, and diagnostic label alignment. All metrics were computed on the 294-note held-out test set and reported as mean ± standard deviation across 5-fold-specific adapters, for both overall and per-class performance across all six glaucoma subtypes. LLM-judge scoring was not performed for GPT-4o due to API access constraints at the time of evaluation.

Statistical comparisons between the fine-tuned Qwen3-8B and zero-shot comparators were performed using two-sided paired t-tests across the five cross-validation folds, with significance set at *P* value < 0.05. Multiple comparisons across the six diagnostic classes were adjusted using the Benjamini–Hochberg false discovery rate correction, with adjusted *P* value < 0.05 considered statistically significant. Subtype-level LLM-judge comparisons between the fine-tuned model and zero-shot Qwen3-8B were additionally assessed using paired t-tests per class. All statistical analyses were performed in Python 3.10 using SciPy v1.13 and statsmodels v0.14.

## Data Availability

All data produced in the present study are available upon reasonable request to the authors

## Data availability

The data that support the findings of this study can be obtained from the corresponding author upon reasonable request.

## Code availability

The codes for the proposed model are available at https://github.com/Mousamoradi/Ci-SSGAN. The fine-tuned Qwen3-8B reasoning model (QLoRA adapters and quantized inference weights) is publicly available on Hugging Face at: https://huggingface.co/spaces/mousamoradi/Reasoning_CISSGAN.

## Acknowledgments

This research was funded by the National Institutes of Health (NIH: R01 EY036222 and R21 EY035298), and MIT-MGB AI Cures Grant. We thank Mass General Brigham’s AI Zone team for providing secure, HIPAA-compliant access to GPT through the Azure AI Foundry platform.

## Author contribution

M.M. conceived the study and contributed to conceptualization, methodology, formal analysis, software, validation, data preparation and cleaning, and writing. A.F. performed data preparation, clinical grading, and review and editing. N.B., D.M.V and K.A. contributed to clinical grading and review and editing. D.L.L. performed clinical grading. M.W. and T.E. provided resources and funding and contributed to review and editing. M.E. contributed to validation, review and editing, project administration, and visualization. N.Z. contributed to clinical grading, review and editing, resources, project administration, and funding.

